# Association of Early Life Risk Factors and APOE ε4 with Incident Dementia: Evidence from 14 Years of Data from the U.S. Health and Retirement Study

**DOI:** 10.1101/2025.04.18.25326086

**Authors:** Eun Young Choi, Gawon Cho, Virginia W. Chang

## Abstract

**Background:** Early life is a critical period for brain development, laying the foundation for cognitive reserve. However, it remains unclear how various aspects of early life relate to dementia risk, and how they may interact with genetic predisposition.

**Methods:** This study leverages data on nationally representative older adults in the United States from the Health and Retirement Study (2006-2020; N=8,676; ages ≥ 60 at baseline, 55% female). We used Cox proportional hazards models to evaluate the associations of early life risk factors (financial, social, human capital deficits, adverse experiences, and childhood health conditions) and their interactions with APOE ε4 genotype on dementia incidence.

**Results:** Low early-life financial, social, and human capital, as well as childhood health conditions were associated with higher dementia risk. After adjusting for comprehensive adulthood risk factors, low social capital and human capital persisted as significant predictors, associated with 16% (95% CI=1–34%) and 21% (95% CI=6–38%) increased risks, respectively. APOE ε4 strongly predicted dementia across all models (83–86% increased risk). Notably, significant interactions emerged between APOE ε4 and early-life financial capital and adversity. These early life risk factors significantly increased dementia risk only among APOE ε4 non-carriers. Carrying ε4, however, consistently elevated risk regardless of childhood socioeconomic status or adversity exposure.

**Conclusions:** Our findings suggest potential direct, enduring cognitive health impacts of inadequate childhood social and human resources, and show that genetic predisposition via APOE ε4 may overwhelm the influence of early life socioeconomic adversity, providing evidence consistent with a differential susceptibility hypothesis.

## Introduction

A life-course approach is increasingly recognized as critical to dementia prevention. Cognitive health in advanced age is a cumulative product of risk factors encountered throughout life [1]. The early years of life constitute a sensitive and crucial period for brain development, laying the foundation for cognitive reserve [2]. Low socioeconomic status (SES) in childhood is found to affect brain maturation patterns, characterized by lower subcortical volume, reduced cortical thickness, and decelerated rates of developmental trajectories [3,4,5]. Individuals growing up in such backgrounds appear to follow different neurobiological development than those from higher childhood SES backgrounds [6]. Thus, exposures to adversities during early life can render certain brain regions more vulnerable to neuropathology and can introduce irreversible alterations in neurotransmitter systems [7].

Recent studies explore how early life experiences influence the risk of dementia in late adulthood. While most studies have focused on SES indicators [8], emerging evidence integrates broader social, psychological, and health-related factors, such as early childhood trauma [9], health conditions [10], and social capital [11]. Early life social relationships, for example, have been linked to cognitive functioning at older ages [12,13,14]. However, many of these studies rely on relatively small, regionally selective samples and cross-sectional cognitive outcomes, highlighting the need for large-scale, longitudinal studies based on a diverse, nationally representative sample of older adults [9,15]. Moreover, early life factors are often studied in isolation from each other, potentially overestimating their independent contributions [16].

Another important yet underexplored area is the interaction between modifiable early life factors and genetic predisposition [7]. The Apolipoprotein E ε4 (APOE ε4) genotype is the single strongest genetic risk factor for dementia, present in 25% of the U.S. population [17]. A handful of studies have investigated APOE ε4-by-childhood interactions, and their findings have been inconsistent. For instance, in South African adults, childhood sexual abuse was associated with memory impairment only among APOE ε4 carriers [18]. Similarly, among older adults in Seattle, low-socioeconomic level paternal occupations were associated with Alzheimer’s disease only for ε4 carriers [19]. In contrast, a study of French adults ages 65+ found a positive childhood environment as a protective factor only for non-carriers [20]. Further, studies from older Australian [21] and Dutch adults [22] reported no interaction effects between childhood adversity and ε4 carrier status for cognitive impairment. This inconsistency underscores a need for further examination of APOE-by-childhood social exposure interactions.

To address these gaps, we examined independent associations between incident dementia and five distinct early life domains (i.e., financial capital, social capital, human capital, adverse experiences, and health conditions), as well as their interactions with APOE ε4. These domains were informed by theoretical frameworks connecting childhood socioeconomic and developmental contexts to cognitive health [10,23,24]. We leveraged 14 years of data from a large, nationally representative cohort to provide robust, generalizable evidence. Current life-course models of dementia prevention primarily highlight lower educational attainment as an early life dementia risk factor [25]. Our comprehensive analysis expands upon this framework, potentially uncovering additional modifiable pathways to dementia and contributing to an evidence-based, lifelong dementia prevention approach.

## Methods

### Data

We used data from the Health and Retirement Study (HRS), a nationally representative cohort of U.S. adults aged 50 and older. In 2006, HRS administered the Psychosocial and Lifestyle Self-Administrated Questionnaire (SAQ), to a randomly selected half of the participants who completed the core interview, with the other half receiving it in 2008. The SAQ collects psychosocial measures used to assess dementia risk factors in our study. Also, in 2006, the HRS began collecting salivary DNA samples, from which APOE genotyping was performed [26]. Therefore, we focused on community-dwelling, dementia-free respondents aged 60+ who completed the SAQ in 2006 or 2008 (n=11,164). Of these, 9,595 had APOE genotyped data. We excluded individuals aged 90+ because they were well past the mean age of dementia onset (n=200) and those missing data on early life factors (n=288), demographics (n=12), or adulthood factors (n=417). Our final sample includes 8,678 participants aged ≥60 and <90 at baseline. Participants were followed through 2020, saving earlier attrition or death without a dementia diagnosis. A sample selection process is presented in **Supplemental Figure 1**.

### Measures

#### Dementia

was identified using the Langa−Weir Classification of Cognitive Function, previously shown to have high sensitivity and good specificity against clinical diagnoses of dementia [27]. Dementia status was determined based on a summary score from the modified Telephone Interview for Cognitive Status. For individuals unable to complete cognitive tests, dementia classification was based on proxy-assessed memory, interviewer-assessed cognitive impairment, and instrumental activities of daily living limitations. Time-to-incident dementia was defined as age midpoint, measured in months, between dementia-free and dementia-classified interviews. Additionally, participants classified as dementia-free under the Langa–Weir classification but later reported in post-death proxy interviews to have been diagnosed with Alzheimer’s disease or other dementias were identified as dementia cases. For these individuals, event time was defined as the age midpoint between dementia-free interview and death. For those who remained dementia-free throughout the study period, event time was defined as either their age at final follow-up assessments or age at death.

#### APOE ε4 isoform

was genotyped through the Taqman allelic discrimination SNP assay, with imputation from existing genotype array data used when direct genotyping was unavailable (12% in our sample) [26]. We used a dichotomized indicator for APOE ε4 carrier status (0 vs. ≥1 allele), as only 2% carried two copies.

#### Early life risk factors

were grouped into five domains: *financial capital, social capital, human capital, adversity,* and *health conditions*, measured retrospectively on respondents’ family background up to age 18. Financial, social, and human capital used measures developed and validated by Vable et al [23]. The financial capital scale assesses financial resources and financial instability. The social capital scale measures family structure and maternal investment. The human capital index is based on parental years of education. Detailed information is available elsewhere [28]. These measures were reverse-coded, where higher scores indicated greater risk. Adversity was assessed using the measure developed by Krause et al [29]. An unweighted sum of four items was calculated: repeating a year of school, trouble with the police, parental substance abuse, and physical abuse by a parent. Health conditions were assessed as an unweighted sum of five indicators: poor overall health, neurological conditions, vision impairment, hearing impairment, and mental health problems. Domain-specific high-risk categories were created using ≥1 SD above mean scores; for adversity and health conditions, this threshold was 1 or lower, thus indicating the presence of any adversity or health condition. Detailed descriptions appear in **Supplemental Table 1**.

#### Adulthood risk factors

included well-established dementia risk domains [25]: *SES* (high school or less education and household wealth bottom quintile); *health conditions* (stroke, heart disease, and diabetes ever told by physicians, hearing problems [self-reported fair/poor hearing or ever using a hearing aid], and obesity [self-reported BMI ≥30]); *health behaviors* (current smoking, heavy alcohol consumption [≥3 drinks/day]; *physical inactivity* [vigorous and moderate activity less than 1/week] [30]); and *psychosocial risk* (depression [8-item CES-D ≥3], high loneliness [3-item UCLA Loneliness ≥6 [31]], and high social isolation [5-item Steptoe Social Isolation index ≥2 [32]]).

#### Demographic covariates

included sex, race/ethnicity, and immigrant status.

### Analyses

We used cause-specific Cox proportional hazards models predicting dementia incidence, using age (in months) as the timescale and adjusting for delayed entry and dementia-free death. The proportional hazards assumption was assessed through visual inspections of Schoenfeld residuals. We first examined associations between early life risk factors and dementia, controlling for demographic covariates. All early-life factors were included simultaneously in a single model (Early Life Only Model). Correlations among these binary indicators of early life risk factors were low to modest (rho range = 0.00–0.24; **Supplemental Figure 2**), indicating minimal concern for multicollinearity. However, given that at least some shared variance exists among the risk factors, explicitly accounting for it by including all risk factors in a single model allowed us to estimate each factor’s independent contribution more accurately. Next, we assessed associations between adulthood risk factors and dementia, again adjusting for basic demographics (Adulthood Only Model). Third, we included both early life and adulthood risk factors in the same model to determine whether the contributions of early life risk factors could be explained by adulthood conditions (Full Model). Finally, we tested whether the associations between early life risk factors and dementia risk varied by APOE ε4 carrier status by adding interaction terms between each early life factor and APOE ε4, while adjusting for the main effects of APOE ε4 and other early life risk factors. To account for the complex survey design, all analyses incorporated sampling weights and adjustments for clustering and stratification, ensuring that our estimates were population-representative. Analyses were conducted using Stata 18.0.

## Results

**Table 1** shows the sample characteristics. Our sample included 8,678 individuals, contributing 78,864 person-years (median follow-up of 10.3 years, range = 0.1–15 years). Over the follow-up period, 1,878 (19.4%, weighted) had incident dementia cases, and 2,752 (29.9%, weighted) died without dementia. In our sample, 25.9% carried at least one copy of the APOE ε4 allele. Regarding early life risk factors, 17.8% were classified as high risk for low financial capital, 15.7% experienced low social capital, 15.8% had low human capital, 31% reported at least one adverse childhood experience, and 25.3% had childhood health conditions.

**Table 1.** Sample Characteristics, the Health and Retirement Study, 2006-2020 (N=8,678)

| Variables | % (Mean ± SD) |
| --- | --- |
| **Dementia Incidence** |  |
| Dementia | 19.4% |
| Dementia-free death | 29.9% |
| Alive without dementia | 50.7% |
| **Genetic Risk Factor** |  |
| APOE ε4 carrier | 25.9% |
| **Early life Risk Factors, High Risk** |  |
| Low financial capital (≥ 1.0, range: -3.0–3.1) | 17.8% |
| Low social capital (≥ 1.3, range: -1.5–5.6) | 15.7% |
| Low human capital (≥ 0.8, range: -2.4–2.6) | 15.8% |
| Any adversity (≥ 1.0, range: 0–4) | 31.0% |
| Any health conditions (≥ 0.9, range: 0–4) | 25.3% |
| **Adulthood Risk Factors, High Risk** |  |
| Low education (high school or below) | 54.9% |
| Low household wealth (bottom quintile) | 19.0% |
| Ever stroke | 7.9% |
| Ever heart disease | 25.4% |
| Ever diabetes | 19.6% |
| Obesity (BMI ≥ 30) | 31.1% |
| Hearing problems | 24.1% |
| Current smoking | 11.4% |
| Excessive drinking (≥ 3 drinks per day) | 6.8% |
| Physical inactivity | 26.0% |
| Depression (≥ 3, range: 0–8) | 18.6% |
| Loneliness (≥ 6, range: 3–9) | 24.0% |
| Social isolation (≥ 2, range: 0–5) | 30.5% |
| **Demographic Covariates** |  |
| Baseline Age (range: 60–89.9) | (71.0 ± 7.9) |
| Sex |  |
| Male | 44.7% |
| Female | 55.3% |
| Race/ethnicity |  |
| Non-Hispanic white | 84.6% |
| Non-Hispanic black | 7.6% |
| Hispanic | 6.1% |
| Non-Hispanic other | 1.8% |
| Immigrant status |  |
| U.S.-born | 92.5% |
| Foreign-born | 7.5% |

*Note.* Estimates were weighted with the survey weights. SD=Standard Deviation. The high-risk group was defined as individuals with values $\geq 1$ SD above the mean. For adversity and health conditions, this threshold was 1 or less, meaning the high-risk group includes individuals with any adversity or health conditions versus none.

**Table 2** displays the risk of dementia associated with early life risk factors. In the Early Life Only Model, low financial capital was associated with a 16% increased dementia risk (95%CI=3–32%). Low social reserve (cause-specific Hazard Ratio [CHR]=1.24, 95%CI=1.06– 1.45), low human capital (CHR=1.46, 95%CI=1.29–1.66), and health conditions (CHR=1.12, 95%CI=1.01–1.25) were also associated with greater risk of dementia. In the Full Model, adjusted for all adulthood risk factors, these associations were attenuated but remained significant for low social capital (CHR=1.16) and low human capital (CHR=1.21). Across all models, APOE ε4 carrier status was associated with an 83 to 86% higher risk of dementia.

**Table 2.** Cox Proportional Hazards Models Predicting Time-to-Dementia Incidence by Genetic, Early Life, and Adulthood Risk Factors.

|  | Early Life Only Model |  |  | Adulthood Only Model |  |  | Full Model |  |  |
| --- | --- | --- | --- | --- | --- | --- | --- | --- | --- |
|  | CHR |  | 95% CI | CHR |  | 95% CI | CHR |  | 95% CI |
| **Genetic Risk Factor** |  |  |  |  |  |  |  |  |  |
| APOE ε4 | 1.83 | *** | (1.63,2.06) | 1.86 | *** | (1.66,2.09) | 1.86 | *** | (1.65,2.09) |
| **Early Life Risk Factors** |  |  |  |  |  |  |  |  |  |
| Low financial capital | 1.16 | * | (1.03,1.32) |  |  |  | 1.11 |  | (0.97,1.28) |
| Low social capital | 1.24 | ** | (1.06,1.45) |  |  |  | 1.16 | * | (1.00,1.34) |
| Low human capital | 1.46 | *** | (1.29,1.66) |  |  |  | 1.21 | ** | (1.07,1.38) |
| Any adversity | 1.11 |  | (0.98,1.26) |  |  |  | 1.04 |  | (0.92,1.17) |
| Any health conditions | 1.12 | * | (1.01,1.25) |  |  |  | 1.06 |  | (0.95,1.18) |
| **Adulthood Risk Factors** |  |  |  |  |  |  |  |  |  |
| Low education |  |  |  | 1.44 | *** | (1.29,1.61) | 1.40 | *** | (1.25,1.56) |
| Low household wealth |  |  |  | 1.47 | *** | (1.27,1.69) | 1.46 | *** | (1.27,1.68) |
| Stroke |  |  |  | 1.77 | *** | (1.53,2.05) | 1.75 | *** | (1.51,2.02) |
| Heart disease |  |  |  | 0.99 |  | (0.89,1.10) | 0.99 |  | (0.89,1.09) |
| Diabetes |  |  |  | 1.26 | ** | (1.09,1.45) | 1.26 | ** | (1.10,1.45) |
| Obesity |  |  |  | 0.81 | *** | (0.71,0.91) | 0.80 | *** | (0.71,0.91) |
| Hearing problems |  |  |  | 1.29 | *** | (1.17,1.42) | 1.27 | *** | (1.16,1.40) |
| Current smoking |  |  |  | 1.28 | ** | (1.07,1.54) | 1.28 | * | (1.06,1.54) |
| Excessive drinking |  |  |  | 0.71 | * | (0.52,0.98) | 0.72 | * | (0.52,1.00) |
| Physical inactivity |  |  |  | 1.19 | * | (1.04,1.36) | 1.19 | * | (1.04,1.36) |
| Depression |  |  |  | 1.38 | *** | (1.20,1.59) | 1.35 | *** | (1.17,1.56) |
| High loneliness |  |  |  | 1.30 | *** | (1.17,1.45) | 1.27 | *** | (1.14,1.41) |
| High social isolation |  |  |  | 1.11 |  | (0.99,1.23) | 1.10 |  | (0.99,1.23) |

*Note.* CHR=Cause-specific Hazard Ratio; All models controlled for basic demographics (i.e., sex, race/ethnicity, and immigrant status), not shown.
\*\*\* $p < .001$ ; \*\* $p < .01$ ; \* $p < .05$ .

In the next model, we tested interactions between APOE ε4 carrier status and each early life risk factor on dementia risk. Significant interaction effects were observed for APOE ε4 × low financial capital (CHR=0.72, 95%CI=0.56–0.95) and APOE ε4 × adversity (CHR=0.67, 95%CI=0.54–0.83). The full suite of estimates for these models are provided in **Supplemental Table 2. Figure 1** shows the estimated CHRs for dementia across groups, using APOE ε4 non-carriers with low early life risk as the reference group. Among non-carriers, individuals with low financial capital had significantly higher dementia risk compared to those with higher financial capital. However, among APOE ε4 carriers, dementia risk did not differ significantly by financial risk. Notably, ε4 carriers without financial risk still had higher dementia risk than non-carriers with financial risk. A similar pattern was observed for adversity. Among non-carriers, having any adverse experiences was significantly associated with a higher dementia risk compared to those with no adverse experiences. However, among APOE ε4 carriers, dementia risk did not significantly differ by adversity. Again, APOE ε4 carriers without adversity had a significantly higher dementia risk compared to non-carriers with adversity.

**Figure 1.**
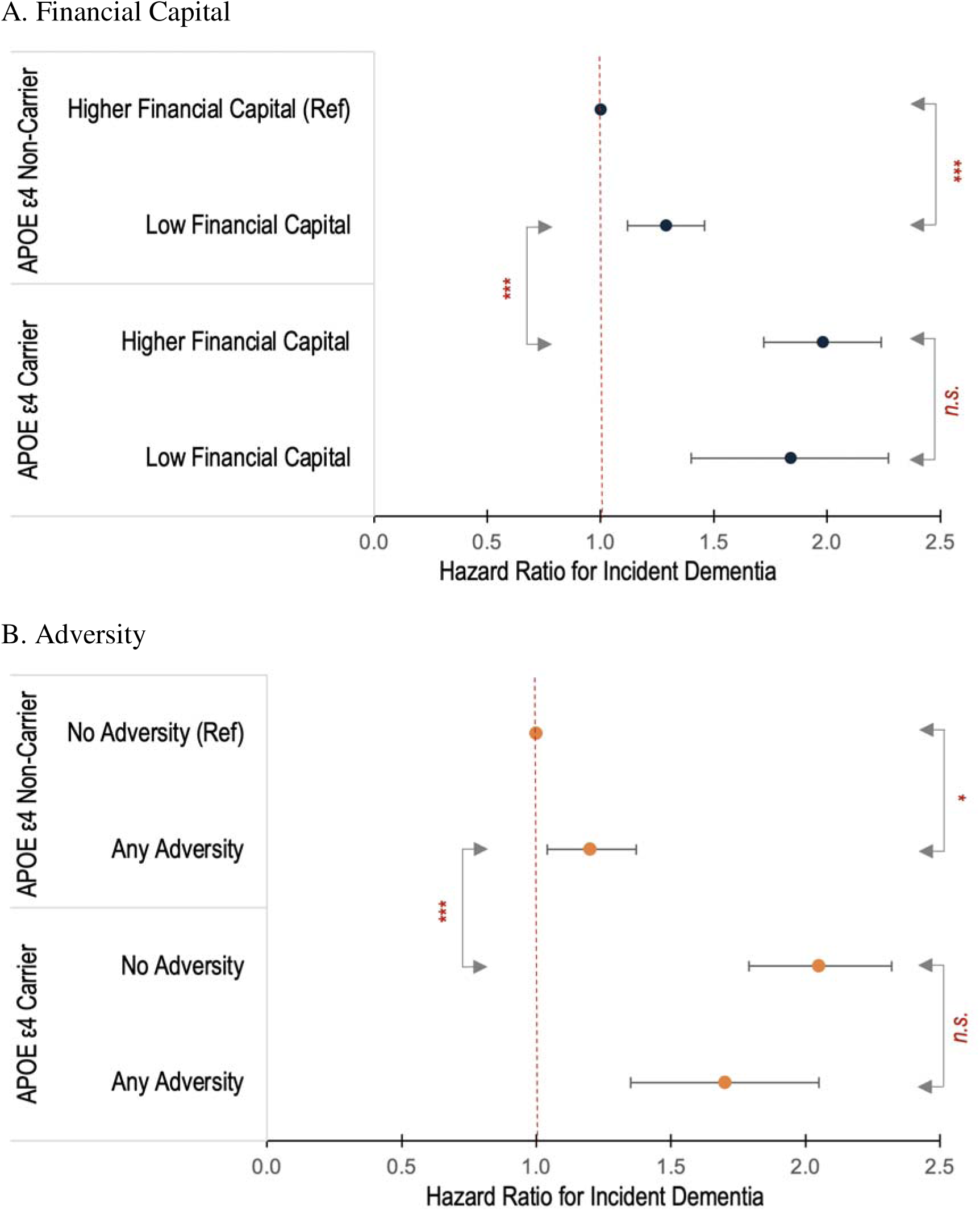
Significant Interaction Effects between APOE ε4 Status and Early Life Risk Factors on Incident Dementia Note. Estimates were adjusted for basic demographics (i.e., sex, race/ethnicity, and immigrant status) and adulthood socioeconomic status (i.e. education and wealth) as well as the main effects of APOE ε4 and all other early life risk factors. *** p < .001; * p < .05. n.s.= Not statistically significant.

We conducted several supplemental analyses to assess the robustness of our findings. First, we repeated analyses excluding APOE ε2 carriers, since the APOE ε2 allele has protective effects against dementia and could potentially confound or attenuate associations observed for APOE ε4. Second, we included an APOE ε4 imputation indicator in the model to account for possible bias introduced by missing genetic data. As shown in **Supplemental Tables 3 and 4**, these additional analyses yielded estimates consistent with our primary findings. Additionally, we conducted supplementary interaction analyses between adulthood risk factors and APOE ε4 status. Significant interaction effects emerged for household wealth and social isolation (**Supplemental Figure 3**). Similar to our findings with early life risk factors, low household wealth and high social isolation significantly increased dementia risk only among APOE ε4 non-carriers, whereas among carriers, dementia risk did not differ significantly by these risk factors.

## Discussion

This study contributes to a growing body of research on early life risk factors for dementia. In previous work, early life risk factors are studied in isolation from each other, which can lead to an inflation in their estimated contribution to dementia risk. In the current study, we simultaneously analyzed multiple early life risk factors in a single model, and found that deficits in financial, social, and human capital, as well as the presence of health problems, are each independently associated with an increased risk of dementia in later life. Furthermore, we observed that adulthood risk factors explained these associations to some degree, suggesting that early life disadvantages may predispose individuals to poorer socioeconomic and lifestyle trajectories in adulthood, thereby increasing dementia risk. Nevertheless, limited social and human capital remained as significant predictors of dementia (16% and 21% increased risks), even after accounting for comprehensive adjustment for adulthood risk factors. Thus, it is possible that these two domains may also directly impact dementia risk by leaving long-lasting effects on neurophysiological and cognitive development *during childhood,* a critical time window that shapes cognitive reserve in later life.

These robust associations may be partially explained by neurodevelopmental changes associated with inadequate social interactions and insufficient cognitive stimulation. Poor parent-child interactions can lead to dysregulated hypothalamic-pituitary-adrenal (HPA) axis activity, chronic inflammation, and increased glucocorticoid levels, which may disrupt neural circuits crucial for cognitive function [33]. Studies using neuroimaging demonstrated that lower parental education is associated with children’s reduced hippocampal volume [34] and thinner prefrontal cortical thickness [35], areas involved in memory formation, executive function, and stress regulation. These structural differences in the brain and physiological disruptions during critical developmental periods may establish biological vulnerabilities that may contribute to neurodegeneration later in life, potentially independently of adulthood conditions.

Importantly, we uncovered significant interactions of the APOE ε4 genotype with early-life low financial capital and adversity. While these risk factors increase dementia risk among ε4 non-carriers, they appear to have little contribution to their already elevated dementia susceptibility for ε4 carriers. Even ε4 carriers without early life disadvantages showed higher dementia risk than non-carriers facing early life social risk. Similar patterns emerged in our supplemental analyses using adulthood socioeconomic and social risk factors, reinforcing the robustness of these genetic-environmental interactions. It is also important to note that the risk factors showing robust independent effects were not the same as those that interacted with APOE ε4, suggesting that certain early life risk factors can have differential influences depending on genetic background and thus, their effects could be obscured if interactions are not considered.

One dominant theoretical framework to explain how APOE interacts with socioenvironmental factors in shaping cognitive health across the life course is the “diathesis stress” model, which argues that individuals carrying APOE ε4 are particularly vulnerable to *adverse* social environments [36,37]. Supporting evidence shows that high social risk in early life disproportionately increases the risk for cognitive declines for APOE ε4 carriers [18,19,38,39].

However, our findings contrast with the diathesis stress model. Early economic disadvantage and adversity was associated with increased dementia risk *only* among APOE ε4 non-carriers. Similarly to our observations, psychosocial stressors, such as depression-evoking environments [40] and loneliness [41], were found to increase memory impairment and dementia risk only for individuals without APOE ε4. Midlife occupational exposure to chemicals or respiratory hazards also increases all-cause dementia risk exclusively in APOE ε4 non-carriers [42]. These findings may be better understood through the “differential susceptibility” hypothesis, which posits that certain genotypes function as plasticity genes, making individuals more responsive to environmental influences [43,44]. In this context, the absence of the ε4 allele may represent a genotype that confers greater sensitivity to environmental conditions. It has been suggested that ε4 carriers have limited neural plasticity and are therefore less responsive to socioenvironmental exposures than non-carriers [37]. ε4 carriers may already be on a more deterministic biological trajectory toward dementia, making external risk factors less influential in altering their risk. APOE ε4 non-carriers, on the contrary, have lower baseline genetic risk, which makes them more responsive to social and environmental influences.

Yet, there remains substantial inconsistency in the literature, with some studies finding minimal or no gene–childhood environment interaction for APOE ε4 [21,22,45,46]. Further work is needed to explain why certain studies, including ours, find ε4 carriers to be less susceptible to childhood social risk factors. One possible explanation for this discrepancy is that genetic influences on cognitive outcomes become stronger with age. Moorman found that the influence of childhood SES on cognition is overwhelmed by the influence of ε4 as people age. Twin and sibling studies also suggests that genetic influences, rather than environmental ones, on cognitive function often intensify with age [37]. Given our sample follows participants to advanced older ages, they may already be at a stage where the genetic influence of APOE ε4 strongly outweighs external impacts. Another consideration is survival bias. We cannot rule out the possibility that individuals carrying APOE ε4 who experienced adverse childhood conditions have lower survival into older age, creating selection effects in older cohorts, and potentially attenuating observed associations.

Several limitations warrant consideration. First, dementia classification relied on cut-off scores established by the Langa-Weir classification, rather than clinical diagnosis, which potentially introduces misclassification bias. Second, we did not distinguish between homozygote and heterozygote APOE4 carriers due to low homozygote prevalence, limiting exploration of dose-dependent genetic effects. Third, early life risk factors were collected retrospectively, relying on self-reported recall of childhood conditions, which may be subject to recall bias, potentially under- or over-estimating true associations.

This study shows the importance of early life socioeconomic and psychosocial environments in shaping later-life dementia risks, emphasizing a clear need for long-term dementia prevention to consider a broader spectrum of early life conditions, particularly those aimed at improving social and human capital. Our findings contribute to the ongoing discussion on gene–environment interactions, suggesting that while childhood socioeconomic risks play a significant role in dementia risk for non-carriers, there can be different pathways of risk accumulation influenced by genetic predisposition. These insights can help refine more tailored dementia prevention strategies that account for both genetic predisposition and early life social conditions.

## Author Approval

All authors have seen and approved the manuscript.

## Data Availability

This study draws on both publicly available data sources and sensitive health data product from the Health and Retirement Study (HRS). Public-use files are freely downloadable from the HRS website at https://hrs.isr.umich.edu. APOE genotype data are part of the HRS Sensitive Health Data Products, which are protected due to privacy and confidentiality requirements. Researchers may apply through the same HRS portal by selecting “Sensitive Health Data Products” and completing the online application.

---

**Supplemental Figure 1.**
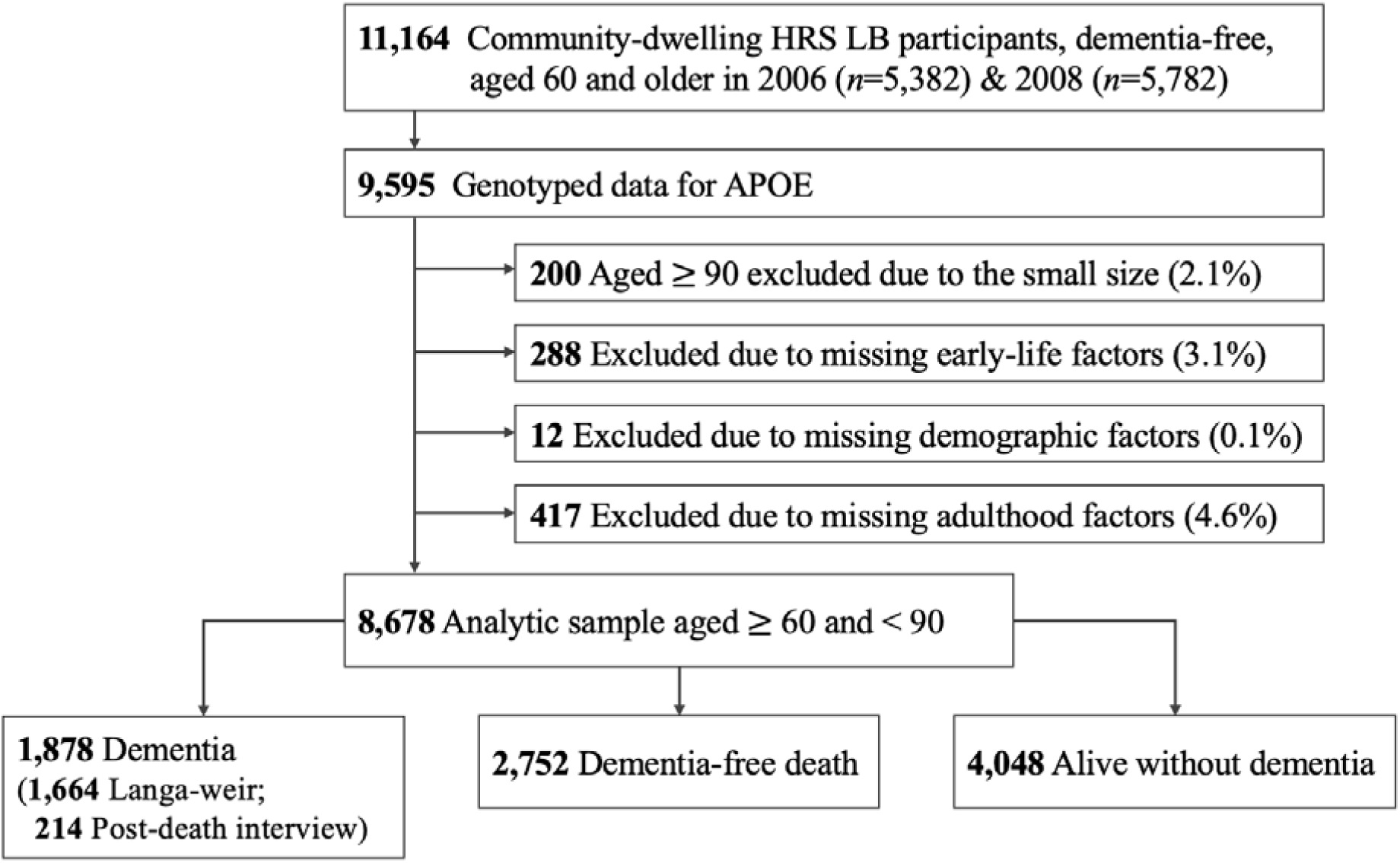
Sample Selection

**Supplemental Figure 2.**
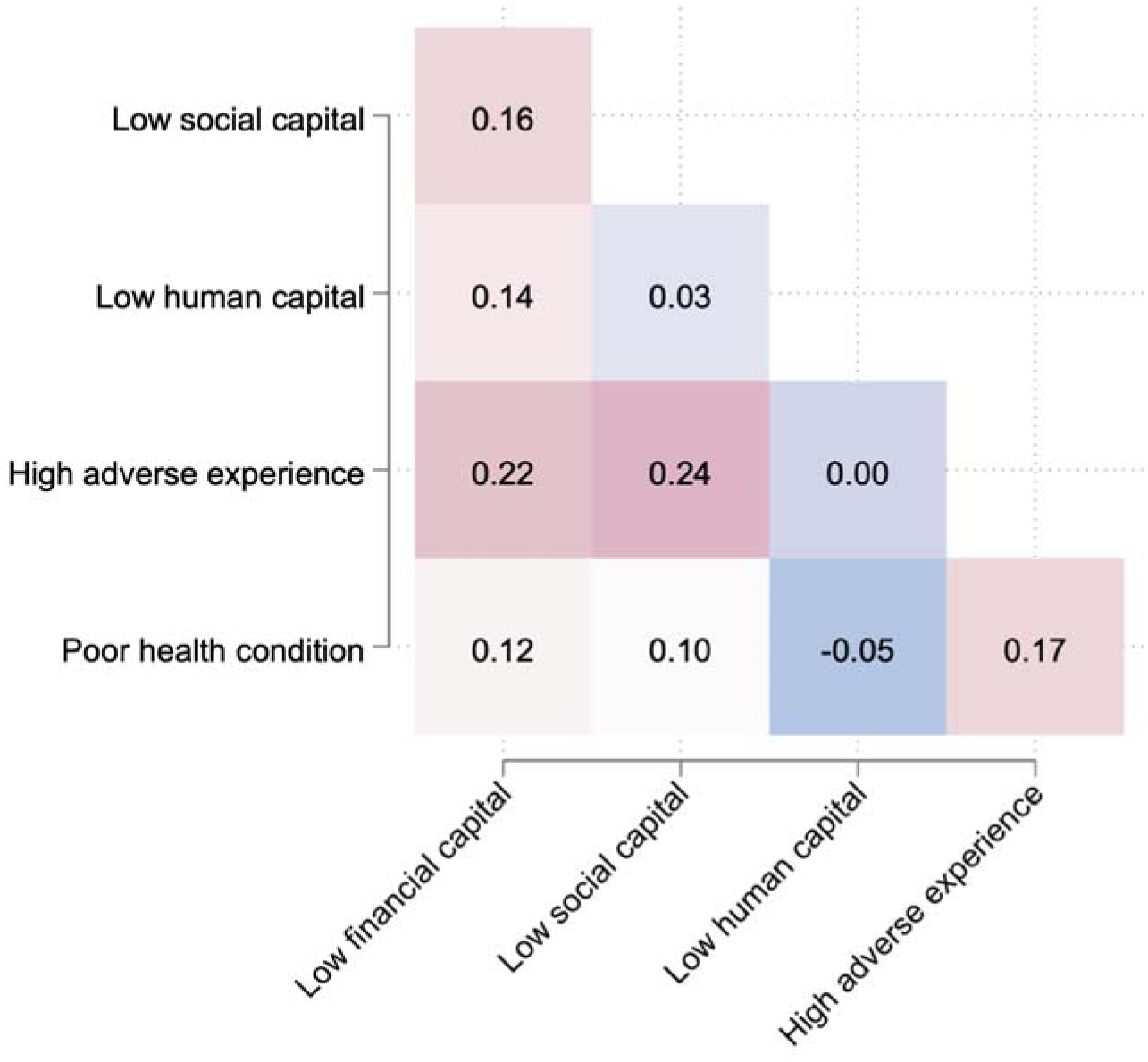
Correlation Among Early Life Risk Factors *Note.* The values represent tetrachoric correlation coefficients (rho)

**Supplemental Figure 3.**
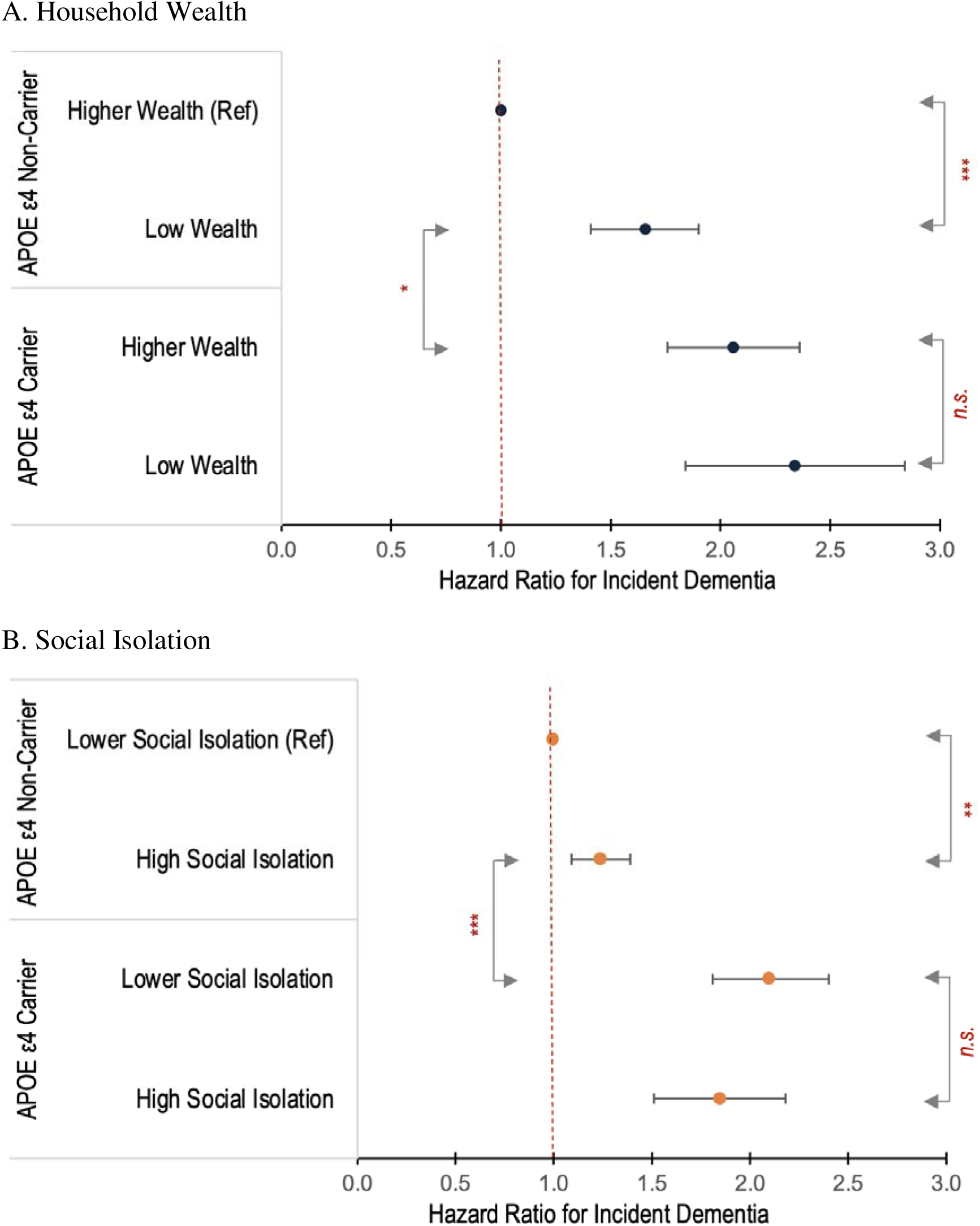
Interactions between APOE ε4 Status and Adulthood Risk Factors and on Incident Dementia Note. Estimates were adjusted for basic demographic characteristics, early life risk factors, and all other adulthood risk factors as well as the main effects of APOE ε4. *** p < .001; ** p < .01; * p <.05.

**Supplemental Table 1.**
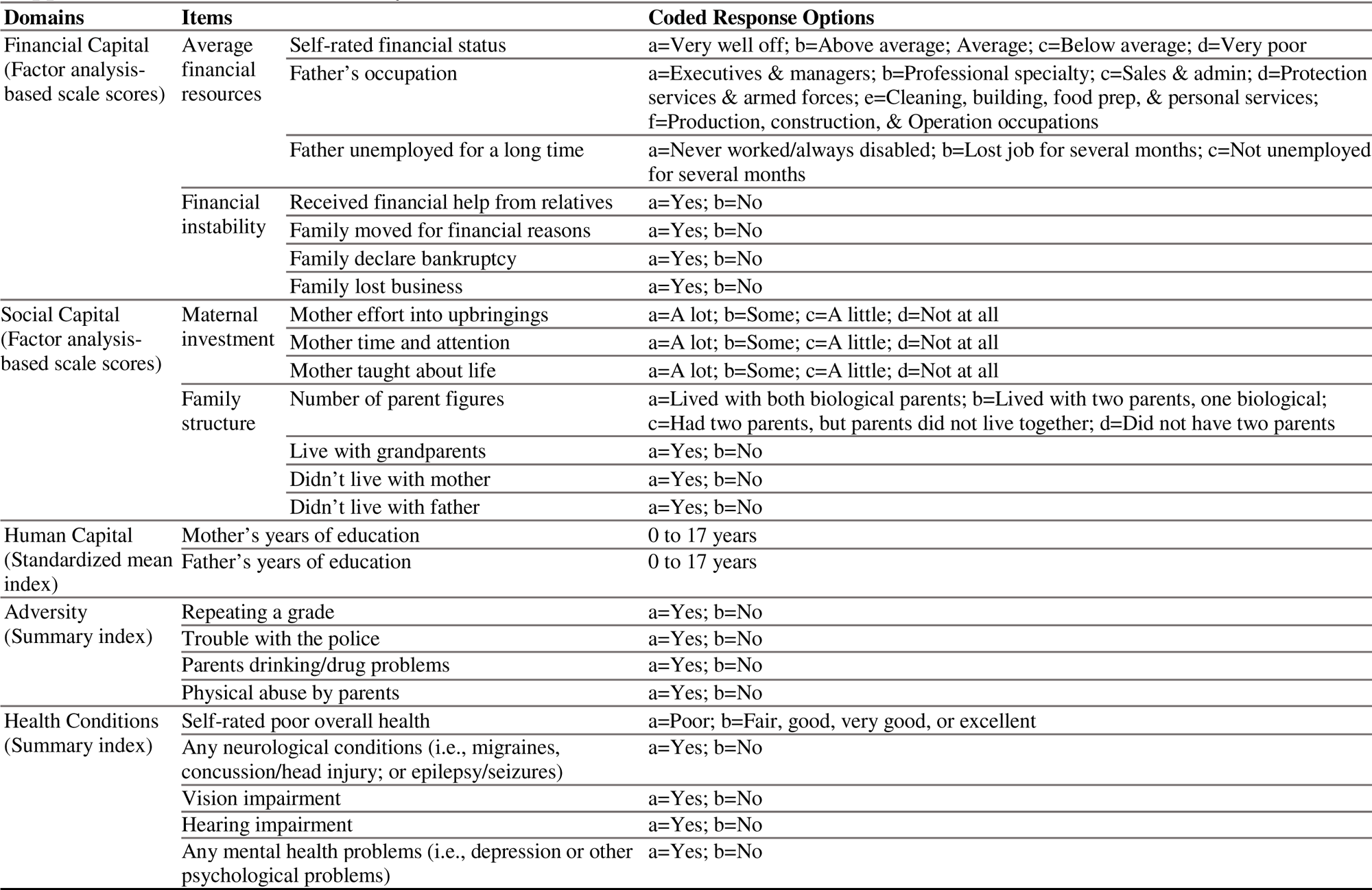
Items for Early Life Risk Factor Domain.

**Supplemental Table 2.** Significant Interaction Effects between APOE ε4 Status and Early Life Risk Factors on Incident Dementia.

|  | Financial Capital Interaction |  |  | Adversity Interaction |  |  |
| --- | --- | --- | --- | --- | --- | --- |
|  | CHR |  | 95% CI | CHR |  | 95% CI |
| **Genetic Risk Factor** |  |  |  |  |  |  |
| APOE ε4 | 1.99 | *** | (1.75,2.27) | 2.09 | *** | (1.82,2.39) |
| **Early Life Risk Factors** |  |  |  |  |  |  |
| Low financial capital | 1.24 | ** | (1.08,1.43) | 1.12 |  | (0.97,1.29) |
| Low social capital | 1.16 | * | (1.00,1.34) | 1.16 | * | (1.00,1.34) |
| Low human capital | 1.21 | ** | (1.07,1.38) | 1.21 | ** | (1.07,1.38) |
| Any adversity | 1.04 |  | (0.92,1.18) | 1.19 | * | (1.03,1.36) |
| Any health conditions | 1.05 |  | (0.95,1.18) | 1.06 |  | (0.95,1.18) |
| **Gene × Early Life Interactions** |  |  |  |  |  |  |
| APOE ε4 × Low financial capital | **0.72** | * | **(0.56,0.95)** |  |  |  |
| APOE ε4 × Any adversity |  |  |  | **0.67** | *** | **(0.54,0.83)** |
| **Adulthood Risk Factors** |  |  |  |  |  |  |
| Low education | 1.40 | *** | (1.26,1.56) | 1.40 | *** | (1.25,1.56) |
| Low household wealth | 1.46 | *** | (1.26,1.68) | 1.46 | *** | (1.27,1.68) |
| Stroke | 1.74 | *** | (1.51,2.01) | 1.75 | *** | (1.51,2.02) |
| Heart disease | 0.99 |  | (0.89,1.10) | 0.99 |  | (0.89,1.10) |
| Diabetes | 1.26 | ** | (1.10,1.45) | 1.26 | ** | (1.09,1.45) |
| Obesity | 0.81 | *** | (0.71,0.91) | 0.80 | *** | (0.71,0.91) |
| Hearing problems | 1.27 | *** | (1.16,1.40) | 1.27 | *** | (1.16,1.39) |
| Current smoking | 1.28 | * | (1.06,1.53) | 1.28 | * | (1.06,1.54) |
| Excessive drinking | 0.72 | * | (0.52,0.99) | 0.72 | * | (0.52,0.99) |
| Physical inactivity | 1.18 | * | (1.03,1.36) | 1.19 | * | (1.04,1.36) |
| Depression | 1.36 | *** | (1.18,1.56) | 1.36 | *** | (1.18,1.57) |
| High loneliness | 1.27 | *** | (1.14,1.41) | 1.28 | *** | (1.14,1.42) |
| High social isolation | 1.11 |  | (0.99,1.23) | 1.11 |  | (0.99,1.23) |

*Note.* CHR=Cause-specific Hazard Ratio; All models controlled for basic demographics (i.e., sex, race/ethnicity, and immigrant status), not shown.
\*\*\* $p < .001$ ; \*\* $p < .01$ ; \* $p < .05$ .

**Supplemental Table 3.**
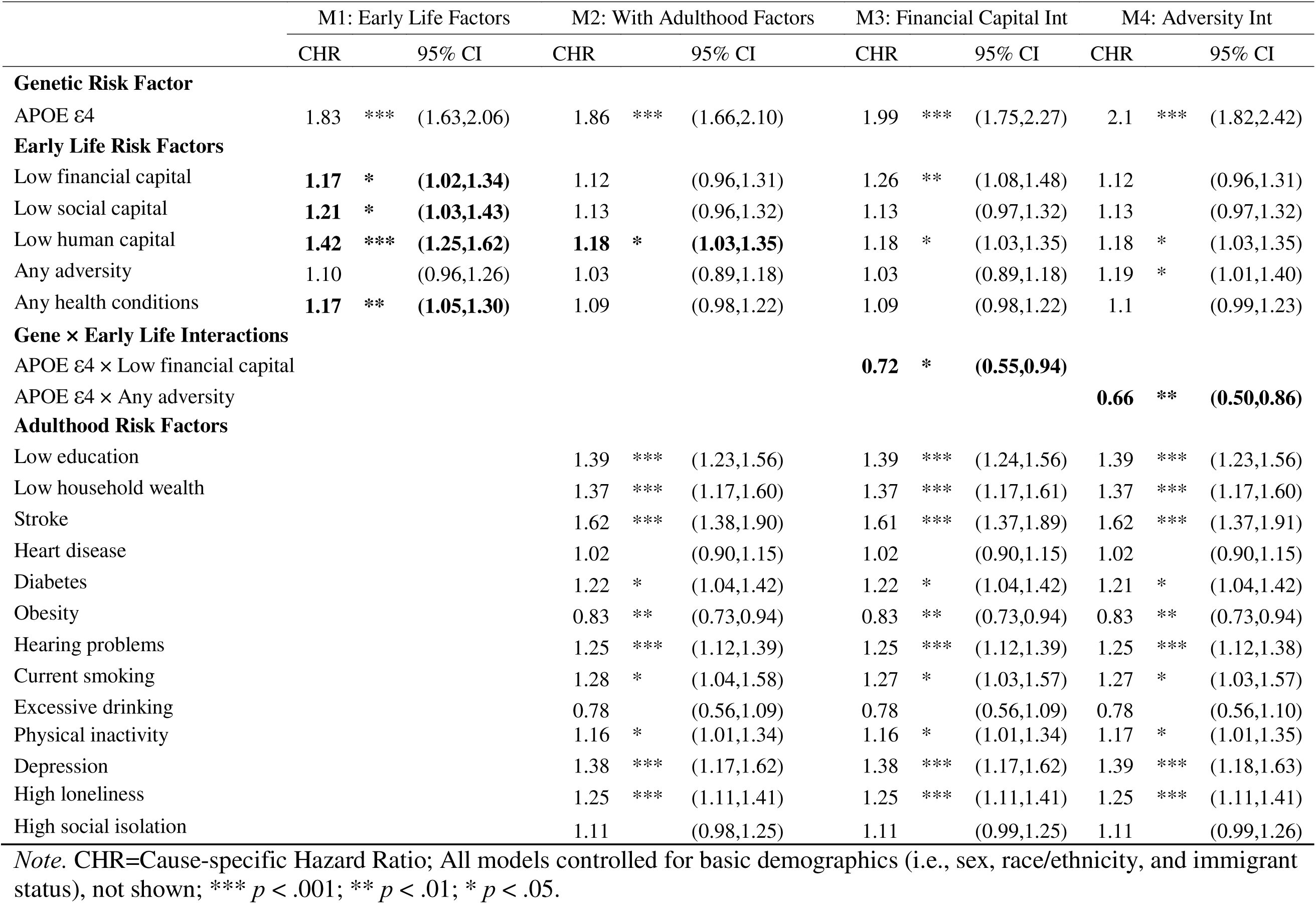
APOE ε4 and Early Life Risk Factors on Incident Dementia, Excluding APOE ε2 Carriers (N=7,355)

|  | M1: Early Life Factors |  |  | M2: With Adulthood Factors |  |  | M3: Financial Capital Int |  |  | M4: Adversity Int |  |  |
| --- | --- | --- | --- | --- | --- | --- | --- | --- | --- | --- | --- | --- |
|  | CHR |  | 95% CI | CHR |  | 95% CI | CHR |  | 95% CI | CHR |  | 95% CI |
| **Genetic Risk Factor** |  |  |  |  |  |  |  |  |  |  |  |  |
| APOE ε4 | 1.83 | *** | (1.63,2.06) | 1.86 | *** | (1.66,2.10) | 1.99 | *** | (1.75,2.27) | 2.1 | *** | (1.82,2.42) |
| **Early Life Risk Factors** |  |  |  |  |  |  |  |  |  |  |  |  |
| Low financial capital | **1.17** | * | **(1.02,1.34)** | 1.12 |  | (0.96,1.31) | 1.26 | ** | (1.08,1.48) | 1.12 |  | (0.96,1.31) |
| Low social capital | **1.21** | * | **(1.03,1.43)** | 1.13 |  | (0.96,1.32) | 1.13 |  | (0.97,1.32) | 1.13 |  | (0.97,1.32) |
| Low human capital | **1.42** | *** | **(1.25,1.62)** | **1.18** | * | **(1.03,1.35)** | 1.18 | * | (1.03,1.35) | 1.18 | * | (1.03,1.35) |
| Any adversity | 1.10 |  | (0.96,1.26) | 1.03 |  | (0.89,1.18) | 1.03 |  | (0.89,1.18) | 1.19 | * | (1.01,1.40) |
| Any health conditions | **1.17** | ** | **(1.05,1.30)** | 1.09 |  | (0.98,1.22) | 1.09 |  | (0.98,1.22) | 1.1 |  | (0.99,1.23) |
| **Gene × Early Life Interactions** |  |  |  |  |  |  |  |  |  |  |  |  |
| APOE ε4 × Low financial capital |  |  |  |  |  |  | **0.72** | * | **(0.55,0.94)** |  |  |  |
| APOE ε4 × Any adversity |  |  |  |  |  |  |  |  |  | **0.66** | ** | **(0.50,0.86)** |
| **Adulthood Risk Factors** |  |  |  |  |  |  |  |  |  |  |  |  |
| Low education |  |  |  | 1.39 | *** | (1.23,1.56) | 1.39 | *** | (1.24,1.56) | 1.39 | *** | (1.23,1.56) |
| Low household wealth |  |  |  | 1.37 | *** | (1.17,1.60) | 1.37 | *** | (1.17,1.61) | 1.37 | *** | (1.17,1.60) |
| Stroke |  |  |  | 1.62 | *** | (1.38,1.90) | 1.61 | *** | (1.37,1.89) | 1.62 | *** | (1.37,1.91) |
| Heart disease |  |  |  | 1.02 |  | (0.90,1.15) | 1.02 |  | (0.90,1.15) | 1.02 |  | (0.90,1.15) |
| Diabetes |  |  |  | 1.22 | * | (1.04,1.42) | 1.22 | * | (1.04,1.42) | 1.21 | * | (1.04,1.42) |
| Obesity |  |  |  | 0.83 | ** | (0.73,0.94) | 0.83 | ** | (0.73,0.94) | 0.83 | ** | (0.73,0.94) |
| Hearing problems |  |  |  | 1.25 | *** | (1.12,1.39) | 1.25 | *** | (1.12,1.39) | 1.25 | *** | (1.12,1.38) |
| Current smoking |  |  |  | 1.28 | * | (1.04,1.58) | 1.27 | * | (1.03,1.57) | 1.27 | * | (1.03,1.57) |
| Excessive drinking |  |  |  | 0.78 |  | (0.56,1.09) | 0.78 |  | (0.56,1.09) | 0.78 |  | (0.56,1.10) |
| Physical inactivity |  |  |  | 1.16 | * | (1.01,1.34) | 1.16 | * | (1.01,1.34) | 1.17 | * | (1.01,1.35) |
| Depression |  |  |  | 1.38 | *** | (1.17,1.62) | 1.38 | *** | (1.17,1.62) | 1.39 | *** | (1.18,1.63) |
| High loneliness |  |  |  | 1.25 | *** | (1.11,1.41) | 1.25 | *** | (1.11,1.41) | 1.25 | *** | (1.11,1.41) |
| High social isolation |  |  |  | 1.11 |  | (0.98,1.25) | 1.11 |  | (0.99,1.25) | 1.11 |  | (0.99,1.26) |

*Note.* CHR=Cause-specific Hazard Ratio; All models controlled for basic demographics (i.e., sex, race/ethnicity, and immigrant status), not shown; \*\*\* $p < .001$ ; \*\* $p < .01$ ; \* $p < .05$ .

**Supplemental Table 4.** APOE ε4 and Early Life Risk Factors on Incident Dementia, Adding APOE Imputation Indicator (N=8,678)

|  | M1: Early Life Factors |  |  | M2: With Adulthood Factors |  |  | M3: Financial Capital Int |  |  | M4: Adversity Int |  |  |
| --- | --- | --- | --- | --- | --- | --- | --- | --- | --- | --- | --- | --- |
|  | CHR |  | 95% CI | CHR |  | 95% CI | CHR |  | 95% CI | CHR |  | 95% CI |
| APOE Imputation Indicator | 1.03 |  | (0.88,1.21) | 1.01 |  | (0.85,1.21) | 1.02 |  | (0.85,1.21) | 1.02 |  | (0.86,1.21) |
| **Genetic Risk Factor** |  |  |  |  |  |  |  |  |  |  |  |  |
| APOE ε4 | 1.84 | *** | (1.63,2.06) | 1.86 | *** | (1.65,2.09) | 1.99 | *** | (1.75,2.27) | 2.09 | *** | (1.82,2.39) |
| **Early Life Risk Factors** |  |  |  |  |  |  |  |  |  |  |  |  |
| Low financial capital | 1.16 | * | (1.03,1.32) | 1.11 |  | (0.97,1.28) | 1.24 | ** | (1.08,1.43) | 1.12 |  | (0.97,1.28) |
| Low social capital | 1.24 | ** | (1.06,1.46) | 1.16 | * | (1.00,1.34) | 1.16 | * | (1.00,1.34) | 1.16 | * | (1.00,1.34) |
| Low human capital | 1.46 | *** | (1.29,1.65) | 1.21 | ** | (1.07,1.38) | 1.21 | ** | (1.07,1.38) | 1.21 | ** | (1.07,1.37) |
| Any adversity | 1.11 |  | (0.98,1.26) | 1.04 |  | (0.92,1.17) | 1.04 |  | (0.92,1.18) | 1.19 | * | (1.03,1.36) |
| Any health conditions | 1.12 | * | (1.01,1.25) | 1.06 |  | (0.95,1.18) | 1.05 |  | (0.94,1.18) | 1.06 |  | (0.95,1.18) |
| **Gene × Early Life Interactions** |  |  |  |  |  |  |  |  |  |  |  |  |
| APOE ε4 × Low financial capital |  |  |  |  |  |  | 0.72 | * | (0.56,0.94) |  |  |  |
| APOE ε4 × Any adversity |  |  |  |  |  |  |  |  |  | 0.67 | *** | (0.54,0.83) |
| **Adulthood Risk Factors** |  |  |  |  |  |  |  |  |  |  |  |  |
| Low education |  |  |  | 1.40 | *** | (1.25,1.56) | 1.40 | *** | (1.26,1.56) | 1.40 | *** | (1.25,1.56) |
| Low household wealth |  |  |  | 1.46 | *** | (1.27,1.68) | 1.46 | *** | (1.27,1.68) | 1.46 | *** | (1.27,1.68) |
| Stroke |  |  |  | 1.75 | *** | (1.51,2.02) | 1.74 | *** | (1.51,2.01) | 1.75 | *** | (1.51,2.02) |
| Heart disease |  |  |  | 0.99 |  | (0.89,1.09) | 0.99 |  | (0.89,1.10) | 0.99 |  | (0.89,1.10) |
| Diabetes |  |  |  | 1.26 | ** | (1.10,1.45) | 1.26 | ** | (1.10,1.45) | 1.26 | ** | (1.09,1.45) |
| Obesity |  |  |  | 0.80 | *** | (0.71,0.91) | 0.81 | *** | (0.71,0.91) | 0.80 | *** | (0.71,0.91) |
| Hearing problems |  |  |  | 1.27 | *** | (1.16,1.39) | 1.27 | *** | (1.16,1.40) | 1.27 | *** | (1.16,1.39) |
| Current smoking |  |  |  | 1.28 | ** | (1.06,1.54) | 1.28 | * | (1.06,1.53) | 1.28 | * | (1.06,1.54) |
| Excessive drinking |  |  |  | 0.72 | * | (0.53,1.00) | 0.72 | * | (0.52,0.99) | 0.72 | * | (0.52,0.99) |
| Physical inactivity |  |  |  | 1.19 | * | (1.04,1.36) | 1.18 | * | (1.03,1.36) | 1.19 | * | (1.04,1.36) |
| Depression |  |  |  | 1.35 | *** | (1.17,1.56) | 1.36 | *** | (1.18,1.56) | 1.36 | *** | (1.18,1.57) |
| High loneliness |  |  |  | 1.27 | *** | (1.14,1.41) | 1.27 | *** | (1.14,1.41) | 1.28 | *** | (1.14,1.42) |
| High social isolation |  |  |  | 1.10 |  | (0.99,1.23) | 1.11 |  | (0.99,1.23) | 1.11 |  | (0.99,1.23) |

*Note.* CHR=Cause-specific Hazard Ratio; All models controlled for basic demographics (i.e., sex, race/ethnicity, and immigrant status), not shown; \*\*\* $p < .001$ ; \*\* $p < .01$ ; \* $p < .05$ .

## References

1. Rodriguez FS. Life-Course Pathways to Cognitive Aging: The Significance of Intellectual Stimulation in the Form of Education and Occupation for Public Policy and Prevention Plans. Frontiers in Psychiatry 2021; 12.

2. Borenstein AR, Copenhaver CI, Mortimer JA. Early-Life Risk Factors for Alzheimer Disease. Alzheimer Disease & Associated Disorders 2006; 20: 63.

3. Vijayakumar N, Allen NB, Youssef G et al. Brain development during adolescence: A mixed-longitudinal investigation of cortical thickness, surface area, and volume. Human Brain Mapping 2016; 37: 2027–2038.

4. Barch DM, Donohue MR, Elsayed NM et al. Early Childhood Socioeconomic Status and Cognitive and Adaptive Outcomes at the Transition to Adulthood: The Mediating Role of Gray Matter Development Across Five Scan Waves. Biological Psychiatry: Cognitive Neuroscience and Neuroimaging 2022; 7: 34–44.

5. Rakesh D, Whittle S. Socioeconomic status and the developing brain – A systematic review of neuroimaging findings in youth. Neuroscience & Biobehavioral Reviews 2021; 130: 379–407.

6. Rakesh D, Whittle S, Sheridan MA, McLaughlin KA. Childhood socioeconomic status and the pace of structural neurodevelopment: accelerated, delayed, or simply different? Trends in Cognitive Sciences 2023; 27: 833–851.

7. Seifan A, Schelke M, Obeng-Aduasare Y, Isaacson R. Early Life Epidemiology of Alzheimer’s Disease – A Critical Review. Neuroepidemiology 2015; 45: 237–254.

8. Korhonen K, Leinonen T, Tarkiainen L, Einiö E, Martikainen P. Childhood socio-economic circumstances and dementia: prospective register-based cohort study of adulthood socio-economic and cardiovascular health mediators. International Journal of Epidemiology 2023; 52: 523–535.

9. Corney KB, West EC, Quirk SE et al. The Relationship Between Adverse Childhood Experiences and Alzheimer’s Disease: A Systematic Review. Frontiers in Aging Neuroscience 2022; 14.

10. Wang G, Zhou Y, Duan J, Kan Q, Cheng Z, Tang S. Effects of adverse childhood health experiences on cognitive function in Chinese middle-aged and older adults: mediating role of depression. BMC Public Health 2023; 23: 1–16.

11. Sachdev PS. Social health, social reserve and dementia. Current Opinion in Psychiatry 2022; 35: 111.

12. Burr JA, Han SH, Peng C. Childhood Friendship Experiences and Cognitive Functioning in Later Life: The Mediating Roles of Adult Social Disconnectedness and Adult Loneliness. The Gerontologist 2020; 60: 1456–1465.

13. Zhang Z, Xu H, Li LW, Liu J, Choi SE. Social Relationships in Early Life and Episodic Memory in Mid- and Late Life. The Journals of Gerontology: Series B 2021; 76: 2121–2130.

14. Lee H, Schafer M. Are Positive Childhood Experiences Linked to Better Cognitive Functioning in Later Life?: Examining the Role of Life Course Pathways. J Aging Health 2021; 33: 217–226.

15. Wang X-J, Xu W, Li J-Q, Cao X-P, Tan L, Yu J-T. Early-Life Risk Factors for Dementia and Cognitive Impairment in Later Life: A Systematic Review and Meta-Analysis. JAD 2019; 67: 221–229.

16. Fanelli D, Ioannidis JPA. US studies may overestimate effect sizes in softer research. Proceedings of the National Academy of Sciences 2013; 110: 15031–15036.

17. National Institute on Aging. Alzheimer’s Disease Genetics Fact Sheet. National Institute on Aging 2023 https://www.nia.nih.gov/health/alzheimers-disease-genetics-fact-sheet (28 September 2023, date last accessed).

18. Savitz J, Van Der Merwe L, Stein DJ, Solms M, Ramesar R. Genotype and Childhood Sexual Trauma Moderate Neurocognitive Performance: A Possible Role for Brain-Derived Neurotrophic Factor and Apolipoprotein E Variants. Biological Psychiatry 2007; 62: 391–399.

19. Moceri VM, Kukull WA, Emanual I et al. Using census data and birth certificates to reconstruct the early-life socioeconomic environment and the relation to the development of Alzheimer’s disease. Epidemiology 2001; 12: 383–389.

20. Ritchie K, Jaussent I, Stewart R et al. Adverse childhood environment and late-life cognitive functioning: Childhood adversity and late-life cognition. Int J Geriatr Psychiatry 2011; 26: 503–510.

21. Lian J, Kiely KM, Callaghan BL, Eramudugolla R, Mortby M, Anstey KJ. No Association Found: Adverse Childhood Experiences and Cognitive Impairment in Older Australian Adults. The Journal of Prevention of Alzheimer’s Disease 2024; 11: 1818–1825.

22. Korten NCM, Penninx BWJH, Pot AM, Deeg DJH, Comijs HC. Adverse Childhood and Recent Negative Life Events: Contrasting Associations With Cognitive Decline in Older Persons. J Geriatr Psychiatry Neurol 2014; 27: 128–138.

23. Vable AM, Gilsanz P, Nguyen TT, Kawachi I, Glymour MM. Validation of a theoretically motivated approach to measuring childhood socioeconomic circumstances in the Health and Retirement Study. PLOS ONE 2017; 12: e0185898.

24. Ding R, He P. Associations between childhood adversities and late-life cognitive function: Potential mechanisms. Social Science & Medicine 2021; 291: 114478.

25. Livingston G, Huntley J, Sommerlad A et al. Dementia prevention, intervention, and care: 2020 report of the Lancet Commission. The Lancet 2020; 396: 413–446.

26. Faul J, Collins S, Smith J, Zhao W, Kardia S, Weir D. APOE and Serotonin Transporter Alleles – Early release. Ann Arbor, MI: Survey Research Center, Institute for Social Research, University of Michigan, 2021.

27. Crimmins EM, Kim JK, Langa KM, Weir DR. Assessment of Cognition Using Surveys and Neuropsychological Assessment: The Health and Retirement Study and the Aging, Demographics, and Memory Study. The Journals of Gerontology Series B: Psychological Sciences and Social Sciences 2011; 66B: i162–i171.

28. Health and Retirement Study. Validated Measures of Childhood Socio-Economic Status. 2018 https://hrsdata.isr.umich.edu/data-products/validated-measures-childhood-socio-economic-status (14 March 2025, date last accessed).

29. Krause N, Shaw BA, Cairney J. A descriptive epidemiology of lifetime trauma and the physical health status of older adults. Psychol Aging 2004; 19: 637–648.

30. Miller MJ, Cenzer I, Barnes DE, Covinsky KE. Physical inactivity in older adults with cognitive impairment without dementia: room for improvement. Aging Clin Exp Res 2022; 34: 837–845.

31. Russell DW. UCLA Loneliness Scale (Version 3): reliability, validity, and factor structure. J Pers Assess 1996; 66: 20–40.

32. Steptoe A, Shankar A, Demakakos P, Wardle J. Social isolation, loneliness, and all-cause mortality in older men and women. Proc Natl Acad Sci USA 2013; 110: 5797–5801.

33. Lupien SJ, McEwen BS, Gunnar MR, Heim C. Effects of stress throughout the lifespan on the brain, behaviour and cognition. Nat Rev Neurosci 2009; 10: 434–445.

34. Noble KG, Houston SM, Brito NH et al. Family income, parental education and brain structure in children and adolescents. Nat Neurosci 2015; 18: 773–778.

35. Lawson GM, Duda JT, Avants BB, Wu J, Farah MJ. Associations between children’s socioeconomic status and prefrontal cortical thickness. Developmental Science 2013; 16: 641–652.

36. Boardman JD, Barnes LL, Wilson RS, Evans DA, de Leon CFM. Social disorder, APOE-E4 genotype, and change in cognitive function among older adults living in Chicago. Social Science & Medicine 2012; 74: 1584–1590.

37. Reynolds CA, Finkel D, Zavala C. Gene by Environment Interplay in Cognitive Aging. In: Behavior Genetics of Cognition Across the Lifespan. Springer, New York, NY, 2014: 169–199.

38. Womersley JS, Spies G, Seedat S, Hemmings SMJ. Childhood trauma interacts with ApoE to influence neurocognitive function in women living with HIV. J Neurovirol 2019; 25: 183–193.

39. Moorman SM, Carr K, Greenfield EA. Childhood socioeconomic status and genetic risk for poorer cognition in later life. Soc Sci Med 2018; 212: 219–226.

40. Reynolds CA, Gatz M, Berg S, Pedersen NL. Genotype–Environment Interactions: Cognitive Aging and Social Factors. Twin Research and Human Genetics 2007; 10: 241–254.

41. Salinas J, Beiser AS, Samra JK et al. Association of Loneliness With 10-Year Dementia Risk and Early Markers of Vulnerability for Neurocognitive Decline. Neurology 2022; 98: e1337–e1348.

42. Williams VJ, Trane R, Sicinski K, Herd P, Engelman M, Asthana S. Midlife and late-life environmental exposures on dementia risk in the Wisconsin Longitudinal Study: The modifying effects of ApoE. Alzheimer’s & Dementia 2024; 20: 8263–8278.

43. Belsky J, Pluess M. Beyond diathesis stress: differential susceptibility to environmental influences. Psychol Bull 2009; 135: 885–908.

44. Ellis BJ, Boyce WT, Belsky J, Bakermans-Kranenburg MJ, van Ijzendoorn MH. Differential susceptibility to the environment: an evolutionary--neurodevelopmental theory. Dev Psychopathol 2011; 23: 7–28.

45. Wang H-X, MacDonald SWS, Dekhtyar S, Fratiglioni L. Association of lifelong exposure to cognitive reserve-enhancing factors with dementia risk: A community-based cohort study. PLOS Medicine 2017; 14: e1002251.

46. Moceri VM, Kukull WA, Emanuel I, van Belle G, Larson EB. Early-life risk factors and the development of Alzheimer’s disease. Neurology 2000; 54: 415–415.

